# Assessing the Safety and Efficacy of DS-7080a Versus Ranibizumab in Retinovascular Disease: A Phase I, Open-Label, Multicenter study

**DOI:** 10.1101/2025.07.07.25330928

**Authors:** Rubbia Afridi, Giorgio Senaldi, Jaclyn Joyce Hwang, Nam Nguyen, Tatsuya Inoue, Brian Berger, Sunil Patel, Zoha Zahid Fazal, Quan Dong Nguyen, Yasir Sepah

## Abstract

**Purpose:** To study the safety and efficacy of DS-7080a in eyes with neovascular age-related macular degeneration (nAMD) and diabetic macular edema (DME) alone or in combination with ranibizumab.

**Design:** This was a dose-escalation-and-expansion study registered under clinicaltrials.gov (NCT02530918) and organized into three parts. Parts 1 (P1) and 2 (P2) involved dose-escalation-and-expansion for nAMD patients while Part 3 (P3) involved dose-expansion-only for DME subjects.

**Subjects:** Patients with nAMD or DME.

**Methods:** In P1, eligible patients were randomized into 3 sequential dose-level cohorts to establish a maximum tolerated dose (MTD) for DS-7080a as 4 mg. The trial thereafter proceeded to P2 where 27 nAMD subjects were randomized to one of the three treatment arms: DS-7080a-4mg-only, ranibizumab-0.5mg-only, or DS-7080a-4mg-plus-ranibizumab-0.5mg. In P3, 18 subjects were randomized to one of the two arms: DS-7080a-4mg or ranibizumab-0.5mg injected thrice. The treatment period was 12 weeks.

**Outcomes:** Primary endpoints were treatment-emergent adverse events (TEAEs) and serious adverse events (SAEs). Secondary endpoints were changes in central retinal thickness (CST) and best corrected visual acuity (BCVA).

**Results:** This study revealed no AE pattern in nAMD or DME subjects receiving DS-7080a. Seven SAEs occurred during the study, four in P2 and three in P3. No SAE in any cohort was drug-related or ocular. 108 TEAEs occurred during the study, 14 in P1, 49 in P2, and 45 in P3. Six were severe TEAEs, three in P2 and three in P3, but none were drug-related or ocular. Six drug-related TEAEs, four of which were ocular (none were severe), occurred during this study, leading to drug discontinuation. No subject died during the study. In nAMD and DME subjects, DS-7080a monotherapy or co-treatment with ranibizumab resulted in no or small improvements in BCVA and CST, while ranibizumab alone resulted in substantial improvements in both metrics.

**Conclusions:** Although safe for eyes with nAMD and DME, DS-7080a does not show efficacy in BCVA improvement and CST decrease when administered alone or in conjunction with ranibizumab. This is inconsistent with non-clinical studies supporting DS-7080a as a therapeutic agent for retinopathy.

This dose-escalation-and-expansion study on the safety and efficacy of DS-7080a shows that although DS-7080a is safe for retinopathy management, it does not improve vision outcomes structurally or functionally when administered alone or concomitantly with ranibizumab.

## Background

Neovascular age-related macular degeneration (nAMD) and diabetic macular edema (DME) are the leading causes of visual impairment and blindness globally^1,2^. While nAMD and DME are distinct entities, angiogenesis and vascular leakage are believed to be crucial in the development of retinopathy in both conditions. The current standard of care for nAMD and DME are intravitreal injections of anti-vascular endothelial growth factor (anti-VEGF) with proven efficacy in clinical trials^3–8^. However, despite their efficacy, the current standard of care presents certain limitations in terms of lack of long-term durability, inconsistent frequency of dosing, and variability in response to therapy. Furthermore, a portion of patients respond suboptimally to anti-VEGF therapies and progression to blindness continues, thus alternative interventions targeting different pathways are needed to combat these diseases^9^.

Roundabout receptors (ROBO) and their slit ligands (SLIT) regulate a number of functions in different cell types including leukocytes, neurons, vascular smooth muscle cells, and endothelial cells^10^. ROBO4 is expressed specifically on endothelial cells, and ROBO4 stimulation has been demonstrated to ameliorate both vascular leakage and angiogenesis in various disease models, including neovascular eye disease^10–14^. Mechanistically, ROBO4 stimulation causes the inhibition of Rac^12^, a Rho family guanosine triphosphatase, which can be activated in endothelial cells by VEGF and other angiogenic factors. Additionally, endothelial ROBO4 expression has been detected in the eyes of patients with neovascular retinopathies, including in the endothelium of choroidal fibrovascular tissue in patients with AMD^15^, indicating that ROBO4 may regulate the development of choroidal neovascularization (CNV) in patients with this condition. Therefore, Rac inhibition may be an effective target to inhibit angiogenesis in patients with retinopathy.

DS-7080a is an agonistic anti-ROBO4 antibody that inhibits Rac via ROBO4 stimulation. Preclinical studies of DS-7080a in primates have not revealed any untoward adverse effects^16^. Additionally, *in vitro* non-clinical studies of DS-7080a have demonstrated to inhibit the migration of human umbilical vein endothelial cells induced by angiogenic factors that had been detected in the eyes of patients with nAMD^15^. *In vivo* studies also illustrated DS-7080a ability block the development of vascular leakage in laser-induced CNV in cynomolgus monkeys^16,17^. Co-administration of DS-7080a plus ranibizumab via intravitreal (IVT) injection in monkeys did not cause any adverse effects due to DS-7080a either^16^. Therefore, we hypothesized that DS-7080a might assist with safely reducing vascular leakage alone or in combination with ranibizumab. The trial was a phase I, first-in-human study designed to evaluate the safety and tolerability of DS-7080a as monotherapy or adjunctive with ranibizumab in patients with nAMD and DME. In addition, we also evaluated the effects of treatments on improving best corrected visual acuity (BCVA) and reducing the central subfoveal thickness (CST) on spectral domain optical coherence tomography (SD-OCT).

## Methods

### Study design

The study was a phase I, first-in-human, multi-center, open-label, multiple dose-escalation-and-expansion clinical trial in subjects with nAMD or DME. The trial was conducted at 12 centers in the United States from July 2015 to January 2018. The trial was divided into 3 parts: part 1 (P1) and part 2 (P2) involved subjects with nAMD receiving dose escalation and dose expansion respectively, while part 3 (P3) involved subjects with DME receiving dose expansion only. P1 consisted of 3 sequential ascending DS-7080a dose levels (1.0, 2.0, 4.0 mg) each comprising of 3-6 subjects following a traditional “3+3” design for a total of 9-18 subjects. The purpose of P1 was to evaluate safety and tolerability of different doses of DS-7080a and establish the maximum tolerated dose (MTD) to be used in P2 and P3. In P2, 27 subjects were enrolled and assigned to one of the following 3 treatment arms: monotherapy with DS-7080a at MTD (4 mg), monotherapy with ranibizumab (0.5 mg), or treatment with both DS-7080a (4 mg) and ranibizumab (0.5 mg). In P3, 20 subjects with DME were assigned to one of the 2 treatment arms: monotherapy with DS-7080a at MTD or monotherapy with ranibizumab (0.3 mg). The study consisted of 8 visits and included a 14-day screening phase, an 84-day treatment phase, and a 28-day follow-up phase. All patients received three treatments at baseline/Day 1, Day 29, and Day 57. The overall study design is illustrated in **Figure 1**.

**Figure 1:**
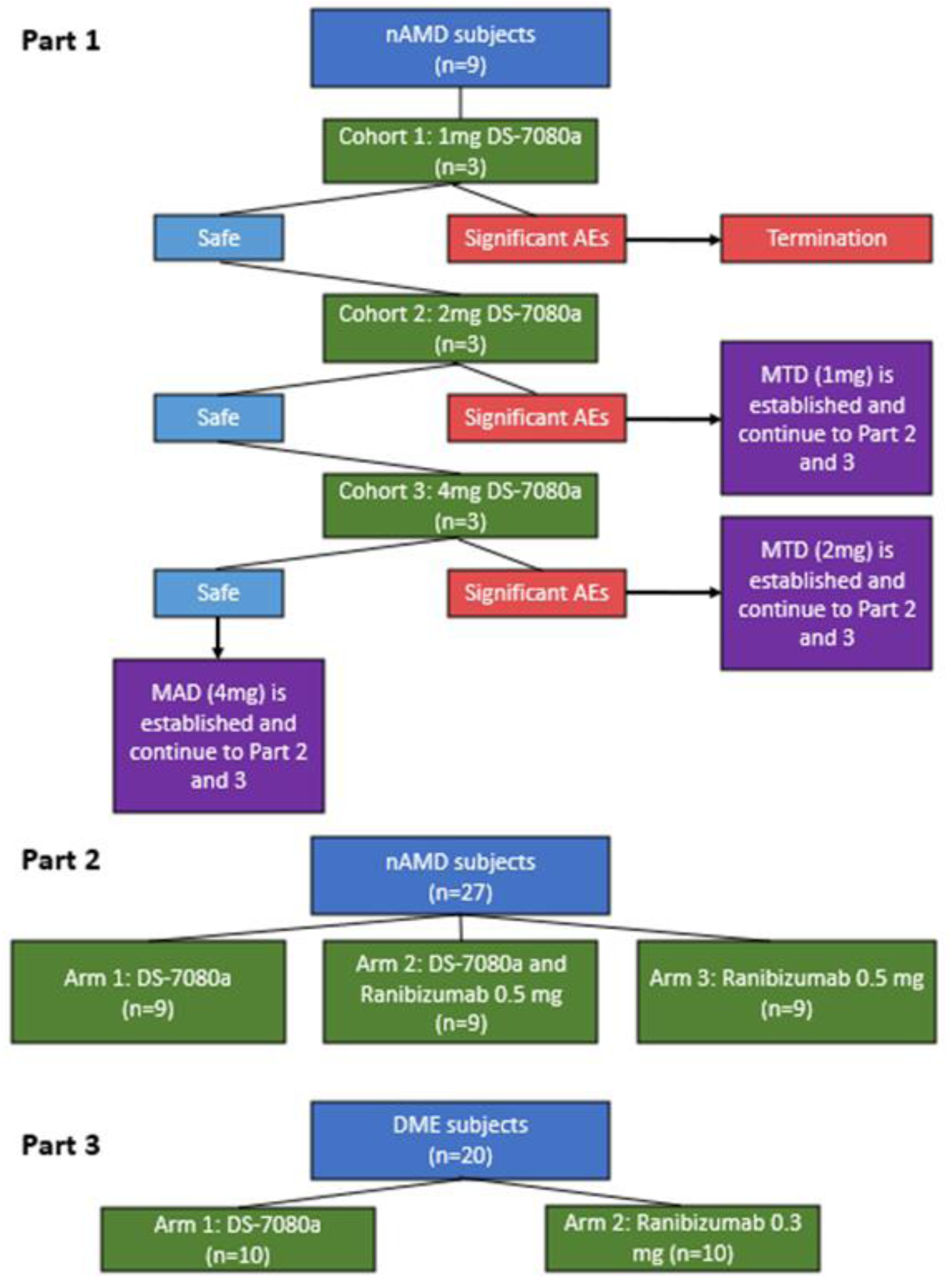
The overall study design and structure for each part of the trial.

### Eligibility Criteria

Participants were eligible for the nAMD cohort if they were ≥ 50 years of age with the following characteristics: active primary sub-foveal CNV lesions secondary to AMD; CNV ≥ 50% of total lesion size in the study eye; CST > 315 µm on SD-OCT as assessed by reading center; and ETDRS BCVA letter score ≤ 49 in the study eye and ≥ 49 in the fellow eye for P1 or ETDRS BCVA letter score of 78 to 25 in the study eye for P2. Participants were eligible for the DME cohort if they were ≥ 18 years of age with the following characteristics: diagnosis of type 1 or type 2 diabetes mellitus and a glycosylated hemoglobin A1c (HbA1c) of ≥ 6.2% and ≤ 12.0%; presence of intraretinal and/or subretinal fluid due to DME with CST of at least 335 µm; leakage observed on fluorescein angiography (FA) in the study eye as confirmed by reading center; and ETDRS BCVA letter score of 78 to 25 in the study eye.

### Ethical Considerations

The study was registered and is publicly available under the <u>clinicaltrials.gov</u> website with study ID NCT02530918^18^. Institutional Review Board (IRB) approval was obtained by the Alpha IRB. The study was conducted in accordance with the CONsolidated Standards of Reporting Trials (CONSORT) guidelines, Declaration of Helsinki, Good Clinical Practice guidelines, and the Health Insurance Portability and Accountability Act. All imaging assessment was performed at the centralized reading center by masked graders. All patients provided informed consent. All patient data was de-identified and stored in password-secure browser only accessible to the study team.

### Study Drug

DS-7080a is a full-length immunoglobulin (IgG2) obtained from mice immunized with the extracellular domain of human recombinant ROBO4. DS-7080a was provided to the site as a frozen sterile solution in a glass vial containing 0.3 mL of an 80 mg/mL DS-7080a solution formulated at pH 5.3 in 10 mM acetate buffer, 100 mg/mL trehalose, and 0.01 w/v% polysorbate 20. It was aseptically diluted with 5% dextrose injection and administered by intravitreal (IVT) injection of 50 µL for each dose (1.0, 2.0, 4.0 mg) in the study eye of each subject. Ranibizumab was administered as intravitreal injections of 0.5 mg (nAMD) or 0.3 mg (DME) ranibizumab aqueous solution containing 10mM histidine HCl, 10% α,α-trehalose dihydrate, 0.01% polysorbate 20 with pH 5.5. An anesthetic following 5% povidone iodine were administered in the study eye before intravitreal injection. In case of combination treatment, ranibizumab was administered at least 30 minutes prior to but no longer than 60 minutes prior to DS-7080a injection. Study drugs were prepared and injected intravitreally according to the instructions from the manufacturer.

### Rescue Criteria

Subjects could be rescued at the discretion of the PI based on the following criteria a) decrease in BCVA in the study eye of 10 or more letters; b) increase of 100 µm in CST (on SD-OCT) due to disease worsening; or c) any reason relating to the health of the subject.

### Assessments

SD-OCT, ETDRS-BCVA and adverse events (AE) query were evaluated at screening, all visits during treatment period (Day 1, 8, 15, 29, and 57), and during follow-up (Day 85 and 113). SD-OCT scans were captured and assessed using Heidelberg Spectralis (Heidelberg Engineering, Heidelberg, Germany). BCVA was assessed according to the ETDRS protocol by certified technicians. AE queries were graded according to the National Cancer Institute common terminology criteria for adverse events (NCI-CTCAE) Version 4.0. Fluorescein angiography (FA) was performed at screening and Day 85, and fundus photography (FP) was captured at Screening, Day 1, 29, 57, 85 and 113. Blood and urine samples were collected at all visits for evaluation of DS-7080a concentration in the system. Physical exam and electrocardiogram were performed at Screening, Day 29, 85 and 113.

### Outcome Parameters

Safety and tolerability of DS-7080a were assessed by monitoring the nature, frequency, and severity of systemic and ocular adverse events, as well as fundus photography findings, vital sign measurements, physical exam and clinical laboratory test results, ECGs recordings, and anti-drug antibody titers. Tolerability was assessed using the NCI-CTAE definitions and subspecifications for treatment-emergent adverse events (TEAE) and serious adverse events (SEA). The efficacy endpoints included: central subfield thickness (CST) on SD-OCT and visual acuity measured in terms of ETDRS BCVA.

### Statistical Methods

No statistical comparisons were made between dose cohorts in P1 of the study, however MTD or the safety and tolerability of MAD was established. Treatment groups within the cohort were compared by exploratory inferential statistics. The statistical summaries were based on the full analysis set (FAS), which include all randomized patients with more than one dose of study drug and more than one measurement of efficacy endpoints. Changes from baseline in CST as measured by SD-OCT and ETDRS BCVA letter score were summarized by treatment and visit, using descriptive statistics including number of subjects (n), mean, standard deviation (SD), median, and range. Comparisons for the mean change from baseline between treatment arms were performed using a repeated measure analysis of covariance (ANCOVA) model with baseline value of the continuous variable and treatment as factors. Ranibizumab treatment was used as the standard reference. The mean treatment difference, standard error, p-value and the 90% confidence interval on the mean difference were presented for each comparison by visit.

## Results

### Baseline Demographics

All enrolled subjects were included in the safety, dose-limiting toxicity (DLT), and FAS data analysis sets. In P1, all subjects were Caucasian adults between ages of 52 and 89 with a mean age (SD) of 67.0 (±7.21), 72.0 (±14.93), and 64.7 (±11.02) years for the 1 mg, 2 mg, and 4 mg treatment groups, respectively. Most subjects in P2 were Caucasian adults between ages of 61 and 90, and mean age between treatment groups were comparable. The safety analysis set for P3 included 20 subjects’ ages 46 to 74 with a mean age of 59.6 (±7.68) years. Out of 36 subjects enrolled in P1 and P2, 35 subjects completed the study and out of 20 enrolled subjects in P3, 17 completed the study. 25 subjects (69%) across treatment groups in P1 and P2 had prior treatment for nAMD. **Table 1** summarizes the patient demographics and baseline characteristics.

**Table 1:**
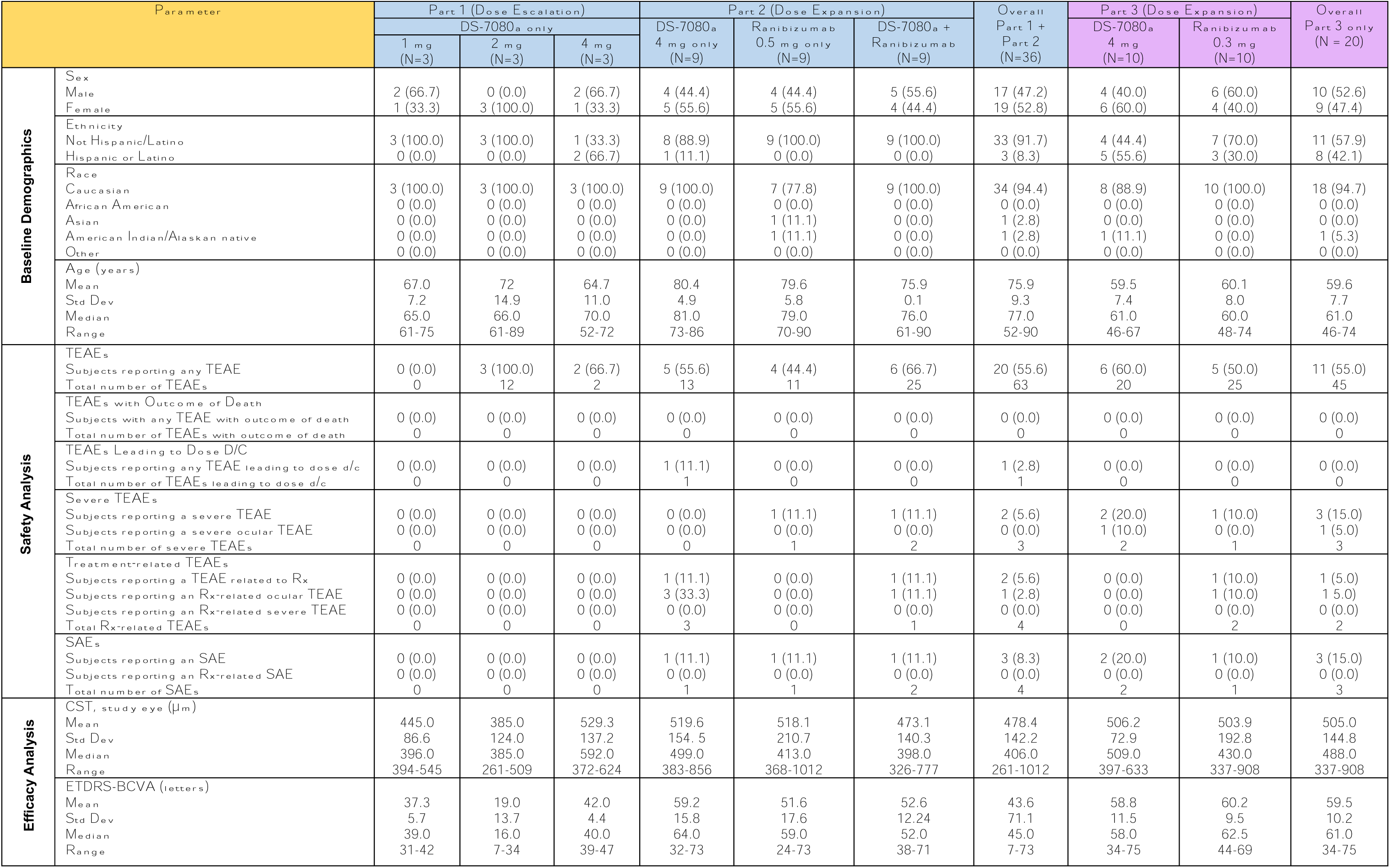
Baseline demographics plus safety and efficacy analyses sets for nAMD and DME subjects in Parts 1, 2, and 3 of the study. All categorical variables (i.e. sex, ethnicity, race, TEAE, and SAE) are reported as n (%). Std Dev: Standard Deviation. d/c: discontinuation. Rx: Treatment.

### Safety Analysis

**Table 1 illustrates** the safety of DS-7080a in patients with nAMD and DME, the details of which are described below. There were no SAEs in any cohort that were related to the study drug. No subject in any cohort died during the study.

#### Treatment-Emergent Adverse Events in nAMD Subjects

In total, 63 TEAEs were reported by 20 subjects (55.6%) in P1 and P2. In P1, 14 TEAEs were reported by 3 (100%) subjects who received 2 mg doses of DS-7080a and 2 (66.7%) subjects who received 4 mg doses of DS-7080a. However, there were no SAEs, no TEAEs rated as severe, and no TEAEs were determined to be related to study drug. 4 mg was thus demonstrated to be safe and well-tolerated in P1, and was established as MAD for P2 and P3.

In P2, 49 TEAEs were reported by 5 (55.6%) subjects in the DS-7080a monotherapy group, 4 (44.4%) subjects in the ranibizumab monotherapy group and 6 (66.7%) subjects in the combination group. Two subjects experienced 3 TEAEs that were rated as severe, but they were non-ocular and not related to the study drug. These include acute mountain sickness in one subject and acute pancreatitis and severe sepsis in the second subject. Two subjects experienced 4 TEAEs that were determined to be related to the study drug. Three of these related TEAEs occurred in one subject in the DS-7080a monotherapy group and consisted in blurred vision, decreased visual acuity (13 letters), and visual distortion which led to the withdrawal of the study drug. Rescue therapy was provided and the subject recovered from this AE. The fourth of these related TEAEs occurred in one subject from the combination group who reported visual distortion (‘seeing black’) for 3 minutes after the second injection (DS-7080a). Additionally, 4 SAEs unrelated to the study drug were experienced by 3 subjects. These consisted of urinary tract infection, acute mountain sickness, acute pancreatitis, and severe sepsis.

#### Treatment-Emergent Adverse Events in DME Subjects

In P3, 11 (55.0%) subjects reported a total of 45 TEAEs; 6 (60.0%) subjects in the DS-7080a group reported 20 TEAEs and 5 (50.0%) subjects in the ranibizumab group reported 25 TEAEs. Three subjects had a TEAE rated severe (2 DS-7080a-treated subjects, 1 ranibizumab-treated subject). These were not related to the study drug. In the DS-7080a treatment group, one subject had increased intraocular pressure and a second had worsening of coronary artery disease. In the ranibizumab treatment group, one subject experienced renal failure. One ranibizumab-treated subject reported 2 TEAEs which were related to the study drug. These were an incidence of subconjunctival hemorrhage and ocular soreness. A total of 2 DS-7080a-treated subjects and 1 ranibizumab-treated subject experienced 3 SAEs. No TEAEs caused a discontinuation of study drug in either treatment group. These TEAEs included urinary tract infection, worsening hypertension, and vitreous hemorrhage.

### Efficacy Analysis

**Table 1** summarizes the changes in BCVA and CST for patients with nAMD and DME patients and have been detailed below. In nAMD and DME subjects, DS-7080a monotherapy or co-treatment with ranibizumab resulted in no or small improvements in BCVA and CST, while ranibizumab alone resulted in substantial improvements in both metrics.

#### BCVA Change in Subjects with nAMD

A total of 27 subjects was enrolled in the P2 of the trial, and one subject from the DS-7080a monotherapy arm received rescue therapy and was discontinued from the study. The baseline BCVA mean of the DS-7080a, ranibizumab, or DS-7080a and ranibizumab treatment groups was 59.2 (15.79), 51.6 (17.59), and 52.6 (12.24) letters, respectively as shown in **Figure 2**. DS-7080a treatment resulted in minimal change in BCVA score from baseline at Day 57 and 113. Treatment with ranibizumab alone resulted in a mean increase in BCVA score of 51.6 (17.59) to 56.6 (17.41) letters from baseline to end-of-treatment visit. Treatment with both DS-7080a and ranibizumab, similar to DS-7080a alone, maintained a near baseline BCVA score. Only at Day 113, BCVA score gain was statistically different in ranibizumab monotherapy group relative to DS-7080a monotherapy group and combination group (DS-7080a treatment group p=0.0074; DS-7080a and ranibizumab group p=0.0249). There was a high amount of variability in individual data. At Day 113, BCVA increased greater than 5 letters in 2 subjects in the DS-7080a monotherapy group.

**Figure 2:**
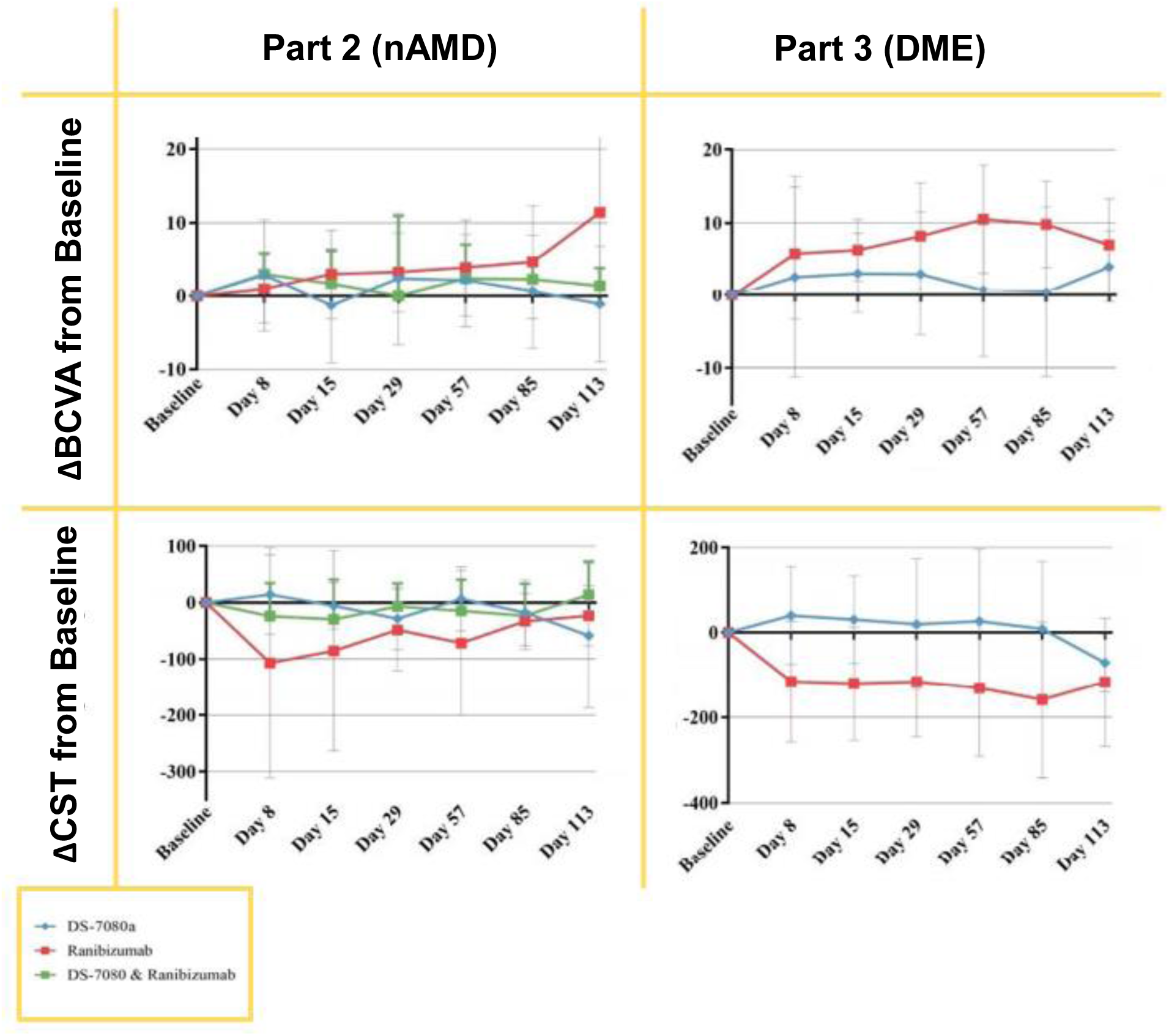
BCVA and CST changes from Baseline in AMD and DME subjects for Parts 2 and 3 of the study.

#### CST Change in Patients with nAMD

The mean CST at baseline in the study eye of the DS-7080a, ranibizumab, and DS-7080a with ranibizumab groups was 519.6 (154.46) µm, 518.1 (210.75) µm, and 473.1 (137.9) µm, respectively. The mean CST change at Day 57 (end of treatment visit) from BL in the study eye of the DS-7080a, ranibizumab, and DS-7080a with ranibizumab groups was 6.4 (65.02) µm, -71.4 (128.84) µm, -14.7 (55.28) µm, respectively and at Day113 was -58.4 (128.22) µm, -23.1 (53.70) µm, and 14.0 (58.80) µm, respectively as shown in **Figure 2**. At Day 8 and Day 57, the DS-7080a treatment group significantly differed from ranibizumab treatment (p = 0.0306, p = 0.0488). A reduction trend was observed in the ranibizumab treatment group relative to the DS-7080a treatment group and the combination after baseline until Day 57. The retinal thickness returned to baseline level in the ranibizumab group 56 days after the last treatment. The retinal thickness in the DS-7080a group did now show any significant changes throughout the study, however the greatest reduction was observed at Day 113.

#### BCVA Change in Subjects with DME

A total of 20 subjects was enrolled in the P3 of the trial, and two subjects in the DS-7080a group dropped out of the study for personal reasons. The baseline BCVA mean (±SD) of the DS-7080a and ranibizumab treatment groups was 58.8 (11.46), and 60.2 (9.46) letters, respectively. The mean BCVA change at Day 57 from BL in the study eye of the DS-7080a and ranibizumab was 0.8 (9.22) letters and 10.5 (7.38) letters, respectively and at Day 113 was 3.96 (2.13) letters and 7.04 (2.00), respectively as shown in **Figure 2**. The greatest increase in mean change from baseline BCVA score in DS-7080a subjects was at Day 113 with 4.08 letters gain, however the visual improvement did not demonstrate a consistent trend throughout the study visits. Treatment with ranibizumab resulted in a mean increase in BCVA score of up to 10.5 letters at Day 57. Only at Day 57, ranibizumab treatment resulted in a significantly different change in BCVA score from baseline relative to DS-7080a treatment (p=0.0275). Visual improvement formed a consistent trend in patients treated with ranibizumab. High variability was observed in both treatment groups at all visits.

#### CST Change in Subjects with DME

The mean (±SD) baseline retinal thickness in the study eye of the DS-7080a and ranibizumab treatment groups was 506.2 (72.95) µm, and 503.9 (192.85) µm, respectively. The mean CST change at Day 57 from BL in the study eye of the DS-7080a and ranibizumab groups was 26.8 (168.75) µm and -130.7 (159.33) µm, respectively and at Day 113 was -71.5 (68.98) µm, and -117.0 (151.11) µm, respectively (Figure 5). Ranibizumab treatment resulted in a statistically significantly greater reduction in retinal thickness compared to subjects treated with DS-7080a from Day 8 to Day 85 of this study at 0.05 levels in pairwise comparison. Ranibizumab treatment maintained a mean of 100 to 200 µm reduction in retinal thickness from baseline. In contrast, the mean change from baseline in the DS-7080a treatment group demonstrated a small increase up until Day 113.

## Discussion

The purpose of this phase I trial was to evaluate the safety and tolerability in patients with nAMD via intravitreal injection of DS-7080a. The drug demonstrated to be safe and was well tolerated in subjects with nAMD up to 4 mg dose. Only two subjects reported TEAEs related to the study drug including blurred vision, decreased visual acuity, and visual distortion. The study also assessed the clinical efficacy of DS-7080a alone or adjunctive to ranibizumab in reducing CST and improving visual acuity. At Day 57 in the nAMD cohort, a statistically significant decrease in CST was observed in the group receiving ranibizumab monotherapy while no significant change was observed in the DS-7080a monotherapy and combination groups. DS-7080a did not show any efficacy as monotherapy or any additional benefits in combination with ranibizumab in term of reducing CST. As established, ranibizumab monotherapy resulted in an overall trend towards improvement in BCVA, however the observed improvement was not statistically significant until Day 113. Meanwhile, DS-7080a did not demonstrate any visual acuity improvement. Additionally, the outcomes in the combination group was less favorable than the ranibizumab alone, which further confirms that DS-7080a is not effective in reducing CST and improving visual acuity in patients with nAMD.

On Day 57 in the DME cohort, a statistically significant improvement in visual acuity and CST reduction was observed in the ranibizumab group, while a similar change was not observed in the DS-7080a group. The efficacy data from two cohorts in this study demonstrated that DS-7080a monotherapy did not provide convincing evidence of improvement in visual acuity and reduction of CST in patients with nAMD and DME. However, the study is a phase I trial, and the sample size is small, therefore any efficacy results warrants cautious interpretation. The data in the study is not consistent with preclinical studies which found DS-7080a blocked the development of vascular leakage in laser-induced CNV, a primate model of nAMD. Additionally, the pharmacokinetic activity of DS-7080a was not measured in aqueous humor or vitreous humor after injection, thus it is not known whether DS-7080a stimulated ROBO4 signaling as it was initially suspected. Future DS-7080a trials need to be conducted carefully before conclusions about DS-7080a clinical efficacy can be made.

Although the study did not meet the secondary endpoints in terms of improvement in BCVA and reduction in CST, some patients in the nAMD cohort showed edema reduction at Day 113 after being treated with DS-7080a alone. Specifically, out of eight subjects enrolled in the DS-7080a monotherapy group, 4 subjects showed fluid reduction at Day 113, in which two subjects showed complete resolution of subretinal fluid, and intraretinal fluid respectively as shown in **Figure 3**. It is not known whether the effects were due to the study drug, however the fluid reduction indicated that the drug might provide some benefits in patients with nAMD.

**Figure 3:**
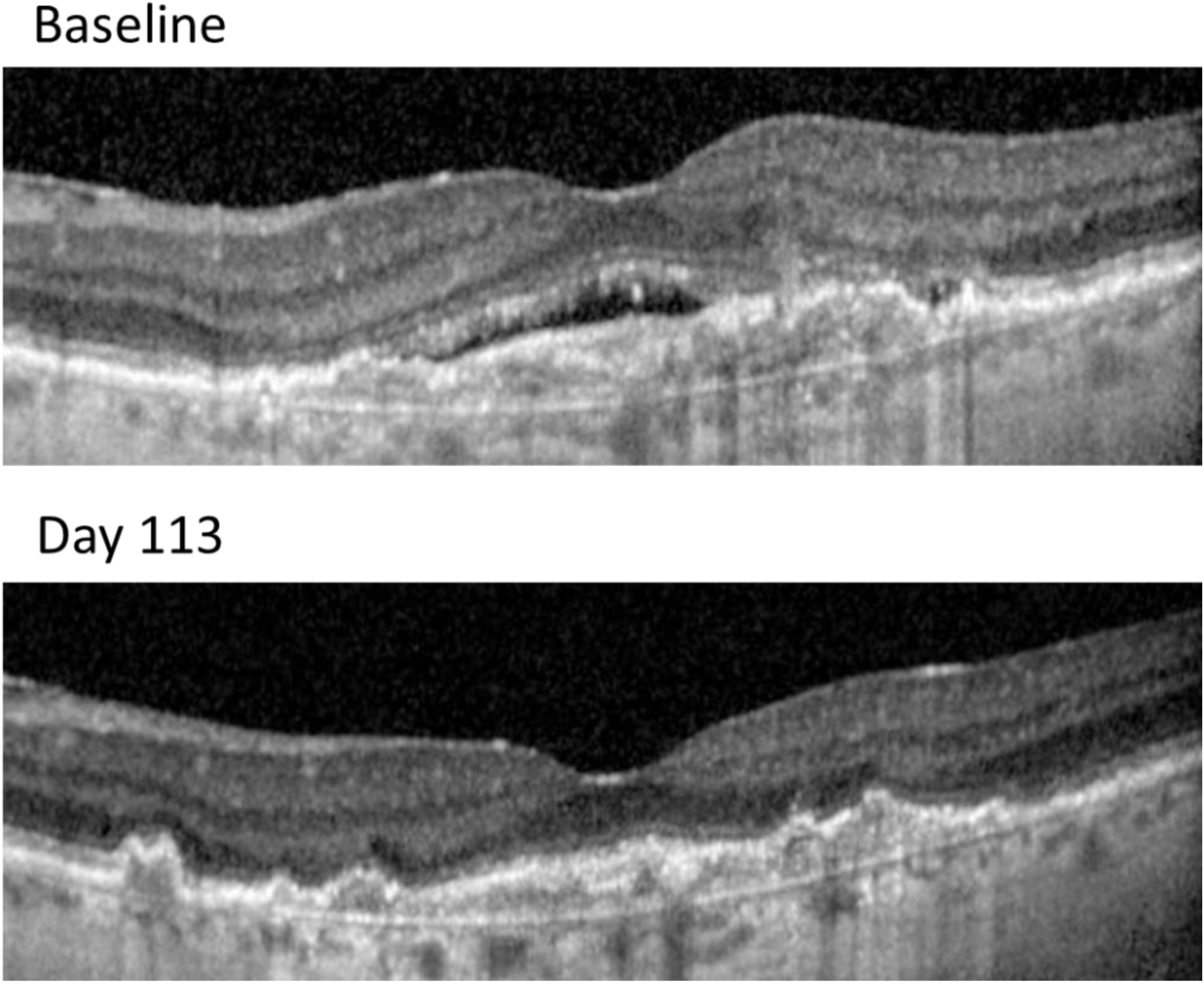
Resolution of subretinal fluid in a subject with neovascular AMD.

This study had several strengths. Patients were randomly assigned to treatments, and all staff at investigative sites and the sponsor were masked to treatment groups. Safety and unexpected adverse events were evaluated and reported by certified staff using the standard protocol. Standardized measures of efficacy were assessed by an independent and experienced reading center. All images were reviewed and assessed by and the reading center using formalized reading protocols. The study also had several limitations. The trial is a phase I study, thus the sample size is small. DS-7080a was hypothesized to decrease macular edema, however central macular edema was not included as an inclusion criterion. Using CST as an efficacy endpoint could not account for edema reduction outside of the central macula. Moreover, DS-7080a protein levels and activity were not measured in the aqueous humor or vitreous humor, therefore whether DS-7080a actually stimulated ROBO4 signaling is not known.

In conclusion, although safe for eyes with nAMD and DME, DS-7080a does not show efficacy in BCVA improvement and CST decrease when administrered alone or in conjunction with ranibizumab.

## Data Availability

All data produced in the present study are available upon reasonable request to the authors.

## Meeting Presentation

The abstract for this manuscript has not been previously presented at any conference.

## Acknowledgments

None.

## Author Contributions

YJS, QDN and GS conceptualized the study and supervised all study processes. RA, BB, and SP collected the data while TI provided preclinical information. YJS and GS analyzed the data. RA, JJH, NN, and ZZF composed the initial draft of the manuscript. All authors reviewed and approved the final submission.

## Abbreviations

AE: adverse event
ANCOVA: analysis of covariance
AMD: age-related macular degeneration
BCVA: best corrected visual acuity
CNV: choroidal neovascularization
CST: central retinal thickness
DLT: dose-limiting toxicity
DME: diabetic macular edema
DR: diabetic retinopathy
DS-7080a: agonistic monoclonal antibody against robo4
ECG: electrocardiogram
ETDRS: early treatment diabetic retinopathy study
FA: fluorescein angiography
FAS: full analysis set
FAZ: foveal avascular zone
FP: fundus photography
HbA1c: hemoglobin A1c
IgG: immunoglobulin g antibody
IRB: institutional review board
IVT: intravitreal
MTD: maximum tolerated dose
nAMD: neovascular age-related macular degeneration
NCI CTCAE: national cancer institute common terminology criteria for adverse events
OCT: optical coherence tomography
OCT-A: optical coherence tomography angiography
P1: first phase of the study
P2: second phase of the study
P3: third phase of the study
ROBO: roundabout protein
Rx: Treatment
SD-OCT: spectral domain optical coherence tomography
SAE: serious adverse event
SLIT: slit ligand for roundabout protein
TEAE: treatment-emergent adverse events
VEGF: vascular endothelial growth factor

